# Qualitative description of interpersonal HIV stigma and motivations for HIV testing among gays, bisexuals, and men who have sex with men in Ghana’s slums - BSGH-005

**DOI:** 10.1101/2023.07.29.23293357

**Authors:** Gamji Rabiu Abu-Ba’are, Edem Yaw Zigah, Osman Wumpini Shamrock, Adedotun Ogunbajo, Henry Delali Dakpui, George Rudolph Kofi Agbemedu, Donte Boyd, Oliver Ezechie, LaRon Nelson, Kwasi Torpey

## Abstract

Despite significant progress in Ghana’s HIV response, disparities in HIV prevalence persist among different populations. Gays, bisexuals, and other men who have sex with men (GBMSM) in the country remain vulnerable to HIV infection due to high levels of stigma and discrimination, limited access to healthcare services, and low HIV knowledge levels. While limited studies focus on HIV prevention and care in the Ghanaian GBMSM context, we did not find studies on GBMSM in slums. We, therefore, explored stigma and motivations of HIV testing among GBMSM in slums. In collaboration with our community partners, we recruited and conducted face-to-face interviews among 12 GBMSM from slums in Accra and Kumasi, Ghana. Our multiple-reviewer summative content analysis identified the following: under HIV stigma, we identified two categories, avoidance of GBMSM living with HIV and fear of testing positive for HIV. Under motivations for HIV testing, we identified three categories; HIV vulnerability, sexual health decision making, and positive messaging about HIV. Our findings provide valuable insights into stigma and motivations for HIV testing among GBMSM in Ghanaian slums. They also highlight the importance of targeted HIV education interventions to empower GBMSM to take responsibility for their sexual health and address the unique challenges they face accessing HIV testing services.

---

Sub-Saharan Africa (SSA) carries over two-thirds of the world’s burden of HIV, yet, HIV testing and related services remain underutilized due to several factors such as insufficient knowledge, low-risk perception, and increased stigma. (1,2) Despite programmatic efforts, the burden of HIV and the factors (e.g., stigma) that hinder access to HIV testing remain a significant obstacle to HIV testing among HIV key populations such as gays, bisexuals, and men who have sex with men (GBMSM) (3–5)

Although Ghana has made significant strides in its HIV response, disparities persist in HIV prevalence between different populations, with GBMSM carrying a disproportionate burden, 18% compared to the nations, 1.7%% (6–8). Efforts to increase HIV testing and prevention among GBMSM continue, yet reports indicate suboptimal testing rates among GBMSM in the country due to individual and environmental barriers (6,7,9,10). At the personal level, such factors include low-risk perception, low HIV knowledge, and fear of rejection (3,11). At the environmental level, factors include healthcare facility-level stigma, inadequate access to testing, and community stigma among others(11–14). At the peer level, interpersonal HIV stigma affects interest in testing as some GBMSM discriminate against their peers living with HIV.(3,11)

While limited studies explore stigma and testing practices among GBMSM in Ghana, no known studies have focused explicitly on GBMSM in Ghanaian slum communities. However, studies in low-income settings identify association between low HIV knowledge and low HIV testing among persons with low socioeconomic status(15,16). Slums also remain associated with high-risk behaviors such as transactional sex, inconsistent condom use, and increased HIV prevalence (17,18). Emerging findings from eastern Africa show that GBMSM in slums have low-risk perception, increased risk behaviors, and low access to HIV testing and prevention services (19,20). For instance, in Kenya, HIV rate among slum residents, 12% was higher than 5% and 6% among non-slum urban and rural residents, respectively (20). Also, out of 4028 youth sampled in Kenyan slums, only 27% had ever tested for HIV, and over 90% had low HIV risk perception. Of those who tested, over 90% reported being required to take the test (19).

Whereas no specific studies exist among GBMSM in Ghana’s slums, previous studies among slum communities in Ghana report increased HIV risk behaviors and higher HIV prevalence among slum residents. These communities also increased HIV prevalence and risk – have poorly resourced infrastructure and health facilities leading to adverse health outcomes (21,22). Urban areas such as the Accra and Kumasi larger regions, where GBMSM were sampled for the study, present the highest prevalence rates of HIV, 2.47% and 1.98%, respectively, compared to the other 16 regions (8). These two cities and their surrounding areas also record high prevalence among GBMSM, Accra (42%) and Kumasi (25%), compared to the national GBMSM rate of 17.5% (6). Thus, placing GBMSM living in urban slums at increased risk of HIV infection than other populations.

The current study seeks to understand GBMSM slum-specific HIV stigma and motivation for testing in Accra and Kumasi, Ghana. Understanding stigma and motivations for HIV testing is critical to reducing HIV transmission and improving health outcomes for this population. As the Self Determination Theory (SDT) explains, individuals’ innate psychological needs inform their well-being and quality of life within a social context (23,24). The various components of SDT (the basic psychological need for autonomy, competence, and relatedness) could help explain the importance of HIV testing motivations among GBMSM in slums. Per the psychological need for autonomy, the barriers faced by GBMSM in slums can undermine the autonomy and restrict their ability to make informed HIV testing (11,12,25,26). On the psychological need for competence, the low levels of HIV knowledge can affect GBMSM’s proficiency to make informed decisions regarding HIV testing (25,27). Also, the stigma and discrimination faced by GBMSM can negatively impact the relatedness aspect of SDT, which emphasizes the need for individuals to feel connected and supported in their social environments. Creating supportive and inclusive environments, reducing stigma, and improving access to healthcare services are crucial for promoting relatedness and encouraging GBMSM to seek HIV testing (28–30).

## Methodology

### Sampling and Recruitment Procedure

Using the time location sampling (TLs) technique, we reached and recruited GBMSM in slum communities in collaboration with our community partners in Accra and Kumasi. Research assistants working with our community partner organizations in Accra (Priorities on Rights and Sexual Health – PORSH) and Kumasi (Youth Alliance on Health and Human Rights – YAHR) screened and invited GBMSM to take part in interviews sessions during one of the organizations’ activities when GBMSM visited the site. We have worked with these community partners in previous studies (9,10). Although we originally intended to find 19 participants, after the eighth interview, we reached saturation and added 4 to ensure complete information saturation, making our total transcripts 12

### Inclusion criteria

Participants in the study had to be at least 18 years old and live in a slum community in Accra, Ghana’s Greater Accra regional capital, or Kumasi, Ghana’s Ashanti regional capital. Additionally, the individual must identify as a cisgender man who self-identifies as gay, bisexual, or pansexual or engage in sexual intercourse with another cis-gender man for reasons other than sexual orientation. The person must have been sexually active during engagement and had sexual intercourse with another cis-gender man within the previous six months.

### Data Collection Procedure

#### Procedure

To gather information from the participants, we conducted in-depth face-to-face interviews. Following the screening, participants were given consent forms by the research assistants to review. The research assistants also read the consent forms out loud and provided extra explanations to ensure everything was understood. They answered the participants’ queries before the interviews started, collected signatures confirming GBMSM’s agreement to participate in the study, and allow for audio recording. The community partners’ private spaces were used for all conversations. All but four interviews were in English, the other four in Twi, a local Ghanaian language that some participants found more conversant. Data were collected from participants from 01/21/2022 to 02/26/2022.

#### Nature of questions

The research assistants were trained to conduct qualitative interviews using the study’s checklist as a guide in collecting information on HIV stigma and motivations for testing among GBMSM living in Ghanaian slum communities. In line with our design, the checklist allowed a more transparent and open discussion rather than the traditional question-and-answer interview structure. Participants were asked to share their experiences of HIV testing, their knowledge, and what motivated or affected their interest in testing.

#### Analytical Strategy

Trained research assistants deidentified the transcripts after translating the audio interview recordings verbatim. We performed a summative content analysis on the transcripts with multiple reviewers (10). Our team has successfully used this analytical method to comprehend crucial components in participant accounts (10). Each transcript received at least two reviewers. Each reviewer independently read the transcripts to identify the most crucial points made by the participants. They then reported these points in between 100 and 200 words. The principal authors reviewed each summary to find clusters and recorded the elements frequently appearing in transcripts and summaries in a data spreadsheet. We identified several clusters and classified them under categories that outlined participant experiences, perceptions, and motivations for HIV testing. Each area that was reported appeared in both peer reviewers’ summaries.

#### Ethical Considerations

Ethical approval was received from the Ghana Health Service Ethics Committee (GHS-ERC 001/10/21) and the Institutional Review Board Committee (IRES IRB) of Yale University (IRES IRB #RNI2000030856). The interviewers in this study ensured that each participant had read and understood the informed consent form thoroughly before any data was collected, and afterward, they obtained written consent.

## Results

### Description of participants

The 12 participants identified as cis-gender men and had sexual intercourse within the previous six months with another cis-gender man. Six participants identified as Christians, four as Muslims, and two as both Muslims and Christians. Five participants accomplished tertiary education, and six concluded senior high school. A member didn’t complete Junior High school.

### Description of findings

We organized the results into two main groups; 1) HIV stigma and 2) Motivations for HIV testing. Under HIV stigma, we identified two categories, avoidance of GBMSM living with HIV and fear of testing positive for HIV. Under motivations for HIV testing, we identified three categories; HIV vulnerability, sexual health decision-making, and positive messaging about HIV.

### HIV Stigma

#### Avoidance of GBMSM living with HIV

Our results show participants had fears and misinformation concerning PLHIV in the slums. Fears stemmed from the possibility of acquiring HIV through everyday activities such as bathing or eating with PLHIV, suggesting a lack of understanding of how HIV is transmitted. Some participants avoided their peers living with HIV or maintained cordial relationships without sexual relations.

> I’m scared of them. I will be scared to eat with the person infected with HIV. Bath with the person or even live in the same area with the person. Because I don’t want to get close to the person, maybe because my thoughts are that the person may transfer it to me at any moment, so I will neglect the person. (GBMSM participant)

> They do much of sex, they are sex addicts. Because when you don’t have sex, you won’t get infected. And married people usually don’t get infected in my community. (GBMSM participant)

> I wouldn’t mind eating with the person because I was told you can only get infected through sex, deep kissing and sharing sharps. So I will treat the person like an ordinary person. Sex will be the only thing I wouldn’t have with this person. I am nondiscriminatory; that’s why my friends came to me and told me they were positive. (GBMSM participant)

#### Fear of testing positive for HIV

Some participants expressed experience testing for HIV. However, they also shared that they are usually scared of testing for HIV due to the fear that they may test positive. Indicating low knowledge of HIV and its treatment options.

> It’s scary, I have tested for HIV, but I won’t like to share my status now. Being in the house, I encourage myself to go and get tested. However, when I get there, it’s scary. What if it comes out positive? But rather, I should be thinking the other way that it should be negative. (GBMSM Participant).

> It’s very scary going for HIV testing, but when you encourage yourself to do the test, you realize it’s not that scary but just like the normal tests that we do. But because of the mentality that if you test, you will come out positive and the ARV and other stuff, its scary. (GBMSM Participant).

#### Motivations for HIV testing among GBMSM

##### HIV vulnerability

Some GBMSM were motivated to test for HIV due to their HIV high-risk awareness, as being a sexually active GBMSM poses a higher risk of getting HIV because of the increased HIV prevalence compared to the general population. Some GBMSM acknowledged that some risky sexual behaviors further increased their risk of acquiring HIV, hence, felt the need to get tested.

> Yes, there’s the need to test for HIV…because men who have sex with men have a higher rate of getting HIV infections compared to men having sex with ladies. So, it’s very good for any MSM to get tested. (GBMSM participant).

> It is very scary when I am going to test. But I feel it’s very important for me to know my status to prevent me from getting sick. So, it’s very important for me to test for HIV. I test for HIV sometimes because maybe I’ve had raw sex with someone, and I don’t trust the person (GBMSM participant).

> I do test for HIV because it’s an opportunistic infection. And you will not know when you might contract it, so I have to test for it to know my status…the sex doesn’t always go as planned…and risky behaviors. I have had some infections some time ago. And now I prefer to be extra careful (GBMSM participant).

###### Sexual health decision making

GBMSM accounts show they are motivated to test for HIV because they want to know their HIV status to make informed decisions about their sexual health. They find testing crucial in seeking appropriate medical care and treatment should they turn positive, thereby promoting long-term health and well-being.

> Of course, it is necessary to get tested because it will help you to know your status and decision-making. if I test positive for HIV, there are medications to help me live long. (GBMSM participants)

> It’s important for me to test for HIV…because you have to know your status. Knowing your status allows you to know what to do. The more you keep on delaying, the more the thing (HIV) too gets worse. Because maybe you are HIV positive, you aren’t on drugs, and if you delay, it might turn into something else (AIDS). And that can cause your death (GBMSM participant).

> I feel it’s important to test for HIV because sometimes you feel like you’ve been exposed (to HIV). Sometimes after having sex, you feel as if you should go, and your body isn’t feeling too well. These thoughts just run through my head, so I go and test because I’m not comfortable with that. So, the earlier you get tested, the better (GBMSM participant).

#### Positive messaging about HIV

Participants cited their knowledge about HIV and the education and sensitization they received about HIV as a motivator for HIV testing. GBMSM in the study recognized that HIV is an opportunistic infection that could affect anyone, and often without any noticeable symptoms until it has progressed to AIDS. As a result, many GBMSM participants emphasized the importance of knowing their HIV status to ensure early detection and access to treatment if needed.

> Lately, I have been hearing a lot about HIV and how it’s spreading. I went and took the test. And I found out I’m negative It is always good to know your status. When I had admitted to college, I had my test before I started lectures. Even if you test positive there are treatment options for you and I learned if you take your medication well, you will achieve the undetectable stage. So, I believe is good to test for HIV (GBMSM Participant).

> I thought HIV was so deadly until I started reading about it. I learned that malaria is even more dangerous than HIV, so I think everyone should test and know their status. And I believe persons living with HIV have the right to life, and they shouldn’t be stigmatized or discriminated against… I’ve learned that there are drugs they give to them when they have HIV. And when they give it to them and take it as prescribed, you will be ok and fine. (GBMSM participant)

> I was told that if I should test positive for HIV, there were treatment options and other medications which would prolong my life. So, I always test when it’s time for me to test. (GBMSM participant)

## Discussion

Despite increased stigma, HIV vulnerability and low HIV testing associated with GBMSM and residents of slums, limited studies examine HIV testing among GBMSM, especially in slums in Ghana (10,31–33). The present study qualitatively describes GBMSM level stigma and their motivations for HIV testing in Ghana’s slums. Whereas some GBMSM demonstrated an understanding and acceptance of people living with HIV, others avoided their peers living with HIV and had fears of testing for HIV. The need to test was driven by factors such as HIV vulnerability, sexual health decision making and positive messaging about HIV-informed motivations for HIV Testing. Highlighting the need for interventions to increase HIV knowledge and leverage the motivation to improve HIV testing among GBMSM in slums (4,34–37).

Although this is one of the early studies to explore GBMSM level HIV stigma in slum communities, the findings remain consistent with literature reported among GBMSM and other populations in Ghana and other SSA (4,34–39). Consistent with previous findings, HIV stigma undermines HIV testing decisions among some participants as they fear testing for HIV due to the stigma associated with testing positive (9,38–40). Thus, it remains imperative for interventions to address HIV stigma to improve testing (9,40,41). Such interventions remain essential considering that the participants did not only have negative misconceptions about HIV, they expressed stigma towards their peers living with HIV by avoiding them, refusing them sex and labeling them as sexually promiscuous. Such labeling and avoidance of others living with HIV reflect our previous findings among other GBMSM in Ghana (9,26,34,40,42). Such behavior poses a significant challenge for testing and the willingness of GBMSM who test positive to disclose their status and even adhere to care as they will not want to be isolated by their peers (42–44).

Despite the stigma shown by others, findings around motivations for HIV testing provide opportunities for leveraging motivated GBMSM in slums to improve HIV knowledge and testing among their peers (10,11,31). As shown in the results, some GBMSM understand the basics of HIV, especially around risk behaviors, and respond to such risk by testing for HIV, which remains consistent with previous research that shows that the awareness of increased vulnerability among GBMSM encouraged them to test to protect themselves and their partners (12,34,45). Consistent with the SDT and prior literature, the participants’ responses also show that increased knowledge of HIV will increase their self-determination for testing. GBMSM did not only consider HIV testing as a path to knowing their HIV status but also as an essential means for knowledge acquisition about their health to enable them to take meaningful steps like seeking care and adopting behavior changes to ensure their well-being. Whereas no previous studies focused on GBMSM in slums, earlier studies among GBMSM in Ghana also showed that some GBMSM acknowledged the importance of HIV testing, as it enabled them to know their status and make important health sexual health behavioral choices (12,34).

To emphasize further, participants’ accounts show that positive messaging about HIV, instead of negative messaging can encourage HIV testing. Others informing participants about how HIV is transmitted and treated, including stages such as undetectable status motivated them to test for HIV. Previous studies reported similar findings among GBMSM (9,38,40). In one of the studies on HIV health promotion, after attending a workshop where GBMSM peer groups discussed and learned about HIV, they observed an increase in HIV testing from 4% to 17% within a week post-intervention (9).

Taken together, the central takeaway from the findings on stigma and motivation for HIV testing lies in the importance of empowering GBMSM to take charge of their sexual health and make informed decisions about HIV testing, as this may help in reducing barriers to testing and promote self-efficacy. By enhancing their sense of competence and autonomy, tailored interventions can motivate GBMSM living in Ghanaian slums to overcome the barriers to HIV testing and take control of their sexual health. A popular intervention that could help reduce HIV stigma at the individual and interpersonal level is the Many Men Many Voices (3MV) (9,40,46). The intervention addresses stigma, HIV risk, transmission, testing, and treatment among GBMSM in Black communities. When adapted to Ghana, we found that GBMSM improved their understanding of HIV, formed community, and improved their HIV testing behaviors (40).

Despite the important findings, research needs to consider the study limitations when applying our participants’ accounts. As a qualitative study, which recruited from two regions in Ghana, the findings may not apply to all GBMSM in slums across other West African regions. We therefore recommend using these results in conjunction with others from the region to draw conclusions. We also recommend using other research designs, like quantitative or mixed methods, to minimize data collection and analysis bias in future studies. Future studies could include GBMSM from other regions and consider targeting specific age groups, as this formative work did not have any specific age brackets and may not fully represent people in different age groups.

In conclusion, our findings contribute to the existing knowledge and provide insights for policymakers, healthcare providers, and researchers to develop effective strategies and programs aimed at reducing HIV disparities and improving HIV testing among GBMSM in slums. While stigma can undermine HIV testing, some GBMSM are highly motivated to test for HIV as such positive messaging about HIV should be encouraged and leveraged to increase HIV self-testing among GBMSM in Ghana’s slums.

## Data Availability

All data contain in the study can be found in the manuscript.

